# On-Campus Dormitories as Viral Transmission Sinks: Phylodynamic Insights into Student Housing Networks During the COVID-19 Pandemic

**DOI:** 10.1101/2025.03.07.25323493

**Authors:** Juan Bolanos, Alex Dornburg, April Harris, Samuel Kunkleman, Jannatul Ferdous, William Taylor, Jessica Schlueter, Cynthia Gibas

## Abstract

University student housing environments are often viewed as hotspots for infectious disease transmission due to their high-density living conditions and high frequency of interpersonal interactions. During the COVID-19 pandemic, concerns arose that on-campus dormitories could serve as amplifiers of viral spread, seeding outbreaks into surrounding off-campus student residences. However, whether on-campus housing acts as a primary driver of transmission or as a recipient of infections introduced from the broader off-campus community remains unresolved. Here, we analyzed 1,431 SARS-CoV-2 genomes collected from students residing on and off campus at the University of North Carolina at Charlotte (UNCC) between September 2020 and May 2022. Using Bayesian phylodynamic and ancestral state reconstruction approaches, we traced viral transmission pathways to determine the directionality of spread between residential settings. Our results indicate that transmission from off-campus housing consistently seeded on-campus dormitory outbreaks. In contrast, viral movement from on-campus to off-campus housing was minimal. These patterns persisted across all major pandemic waves, regardless of shifting mitigation strategies, and suggest that on-campus residences acted as transmission sinks rather than sources of broader student outbreaks. These findings raise the possibility that on-campus residences may be more vulnerable than often considered, functioning as epidemiological ‘islands’ that primarily receive infections from off-campus sources.

**Author Summary:** University dormitories are often considered high-risk environments for disease spread due to their crowded living conditions and the inter-connectivity of individuals. This risk has led to the perception that university student housing could act as a major driver of outbreaks, raising concerns that on-campus dormitories fuel transmission into surrounding off-campus residences and the broader community. However, the actual transmission dynamics between on and off campus communities are often unclear. To address this, we analyzed over 1,400 SARS-CoV-2 genomes from students living on and off campus at the University of North Carolina at Charlotte between September 2020 and May 2022. Using Bayesian phylodynamic and ancestral state reconstruction approaches, we mapped viral transmission pathways and found that infections consistently flowed from off-campus residences into on-campus dormitories—not the other way around. This pattern persisted across multiple waves of the pandemic, regardless of changing mitigation strategies such as masking, testing, and vaccination policies. Our findings suggest on-campus housing functioned more like transmission “islands”—receiving infections from off-campus networks but rarely exporting them. These results suggest that universities should shift their focus toward managing transmission in off-campus student communities rather than concentrating mitigation efforts solely on dormitory settings. By reframing on-campus housing as a transmission *sink* rather than a *source*, our study provides critical insights for improving future outbreak and pandemic response strategies in university environments.

## Introduction

The COVID-19 pandemic has underscored the critical importance of understanding viral transmission dynamics in densely populated environments. University campuses, which serve as microcosms of broader societal interactions, provide a unique lens for studying the spread and evolution of infectious diseases [1]. These environments are characterized by high mobility [2], tightly connected contact networks [3], and varied mitigation strategies in the face of disease outbreaks [4], making them ideal models for understanding how interventions shape transmission within and between residential settings. However, the relationship between on-campus and off-campus transmission is complex and context-dependent. Some studies suggest that reopening campuses can significantly elevate transmission rates in surrounding communities [5,6] with estimates indicating potential increases of over 50% [7]. Conversely, robust on-campus interventions such as testing, contact tracing, and wastewater surveillance can successfully limit spillover between residential groups [8–10]. These heterogeneous outcomes highlight the need for nuanced, data-driven analyses of campus-community transmission patterns to inform context-specific policies to optimize public health outcomes.

Mathematical models and empirical datasets analyzing COVID-19 transmission have identified University campuses as possible superspreaders [11]. These findings are supported by additional studies demonstrating that university-related gatherings and close-contact environments played a significant role in local outbreaks during peak transmission periods [12]. However, the extent to which on-campus dormitories contribute to spread across dispersed off-campus university residential settings is not clear. The dynamics of dormitory populations may differ from the dynamics of the university population as a whole. For example, some universities implemented intervention strategies that significantly lowered transmission within campus housing communities [13]. These outcomes align with mathematical simulations that suggest robust contact tracing and demonstrate that intervention programs can successfully mitigate outbreaks in university settings [14]. College campuses also vary dramatically in their demographics and geographical settings, both of which are highly correlated to incidence patterns [15–17]. University residential settings span the full urban to rural gradient, from campuses located within major population centers to those located within small isolated communities. Most studies have focused on isolated extremes of this spectrum of settings [12], raising the question of how universities in urban environments with off-campus residences scattered throughout high density metropolitan housing areas differ in transmission risks and dynamics.

Urban campuses are characterized by high population densities, extensive student clustering, and substantial daily movement between on campus dormitories, off-campus apartments, and other communal spaces [18,19]. The high frequency of interpersonal interactions, shared spaces, and proximity to common gathering areas could position on-campus housing as viral incidence amplification sites that facilitate streams of transmission to off-campus residences. An alternative hypothesis, though, is that on-campus residences may act as epidemiological ‘islands’, where infections are introduced by contact with individuals residing off-campus who have had greater exposure to contacts in the broader community. In this scenario,viral strains are introduced to campus, and may circulate within the campus population, but contribute little to overall community spread beyond the campus [8,20]. Supporting this latter scenario, Purdue University detected at least 10 independent introductions of the gamma variant onto its campus in 2021 [21], emphasizing the possibility of multiple entry points into on campus residences. Disentangling these contrasting hypotheses–akin to classic source-sink dynamics in biogeography [22–25]– is critical for understanding the degree to which student housing acts as a self-contained transmission network or as a gateway for broader community spillover.

To address this question, we apply a methodological approach capable of tracing viral movement with high resolution. Traditional epidemiological metrics such as incidence data have been widely used to reveal overall trends in infection rates between campus and other environments [5]. However, these metrics cannot resolve the history of transmission between individuals in the absence of detailed contact tracing. Recent advances in phylodynamic approaches facilitate the integration of viral genomic data with epidemiological models permit reconstruction of transmission pathways with robust resolution [26,27]. Such approaches have been widely used to track the spread of infectious diseases at global and national scales [28–30]. Instances of their application to university campuses is less common [12], creating a knowledge gap in our understanding of general trends in the transmission dynamics of viruses between campus populations.

Here we integrated extensive SARS-CoV-2 genomic sequence data (n=1431 genomes) with public health and intervention data from the University of North Carolina at Charlotte (UNCC), a mid-sized urban institution serving over 31,000 students within the densely populated Charlotte metropolitan region [31]. We analyzed these genomic and epidemiological data in a Bayesian phylodynamic framework. We apply time-calibrated phylogenies and discrete ancestral state reconstructions [32] to test three key hypotheses regarding the role of student housing settings in transmission dynamics. First, we assessed the degree to which transmission dynamics between on- and off-campus populations reflect alternate possible source-sink scenarios. Specifically, we determined whether the university primarily drives spillover into surrounding off-campus housing or functions as a recipient of infections introduced from off-campus residences. Second, we evaluated whether shifts in on-campus mitigation strategies, such as masking mandates, asymptomatic and symptomatic testing, and dormitory occupancy limits, corresponded to measurable changes in transmission patterns within and between campus and off-campus residences. Lastly, we examined whether viral lineage dynamics in residences represented early indicators of broader outbreaks or if infections lagged behind those in the greater Charlotte metropolitan area. Collectively, our results challenge the prevailing popular media notion that on campus residences act as primary superspreader hubs or reservoirs amplifying off-campus transmission. Instead, they show a higher rate of transmission into the campus under all mitigation conditions examined.

## Results & Discussion

### Campuses dormitories can act as sinks of viral transmission

We sequenced viral genomes from 1444 SARS-CoV-2 positive clinical samples collected at the University of North Carolina at Charlotte (UNCC) between September, 2020 and May, 2022. These samples were from 470 and 961 patients living on-campus or off-campus, respectively. Using this data, we estimated a time calibrated Bayesian phylogenetic framework to model the transmission pathways of SARS-CoV-2 between on-campus and off-campus student populations via ancestral state reconstruction (ASR).(**Figure 1A**). Given the location of the school and the large number of commuter students, our initial hypothesis was that on campus dormitories could act as a primary source of infections, fueling outbreaks among off-campus students and facilitating spread into the broader community. However, our results strongly support that this is not the case. (**Figure 1**). Assessing the distribution of off and on campus incidence across the phylogenetic history of transmission revealed numerous shifts between on and off campus residences (**Figure 1A**). In most cases, transmission pathways reflect bursts of infections within either the on or off campus community, reflecting a higher rate of transmission within either community than between communities (**Figure 1B**). In cases of transmission between on and off campus residences, transmission from off-campus residences into campus residences dramatically outnumber the reverse (**Figure 1C).** This asymmetry suggests that viral introductions into the university environment were primarily seeded from off-campus sources rather than spreading outward into the community, with as many as one fifth of the total number of cases on-campus being a direct consequence of transmission from off-campus residents. These results were robust to bootstrapping of the data (**Supplemental Figure 2**), and they challenge assumptions that university dormitory populations act as amplifiers of community transmission. Rather, our findings suggest the on-campus student population is a transmission sink within the larger context of the student housing network.

**Figure 1.**
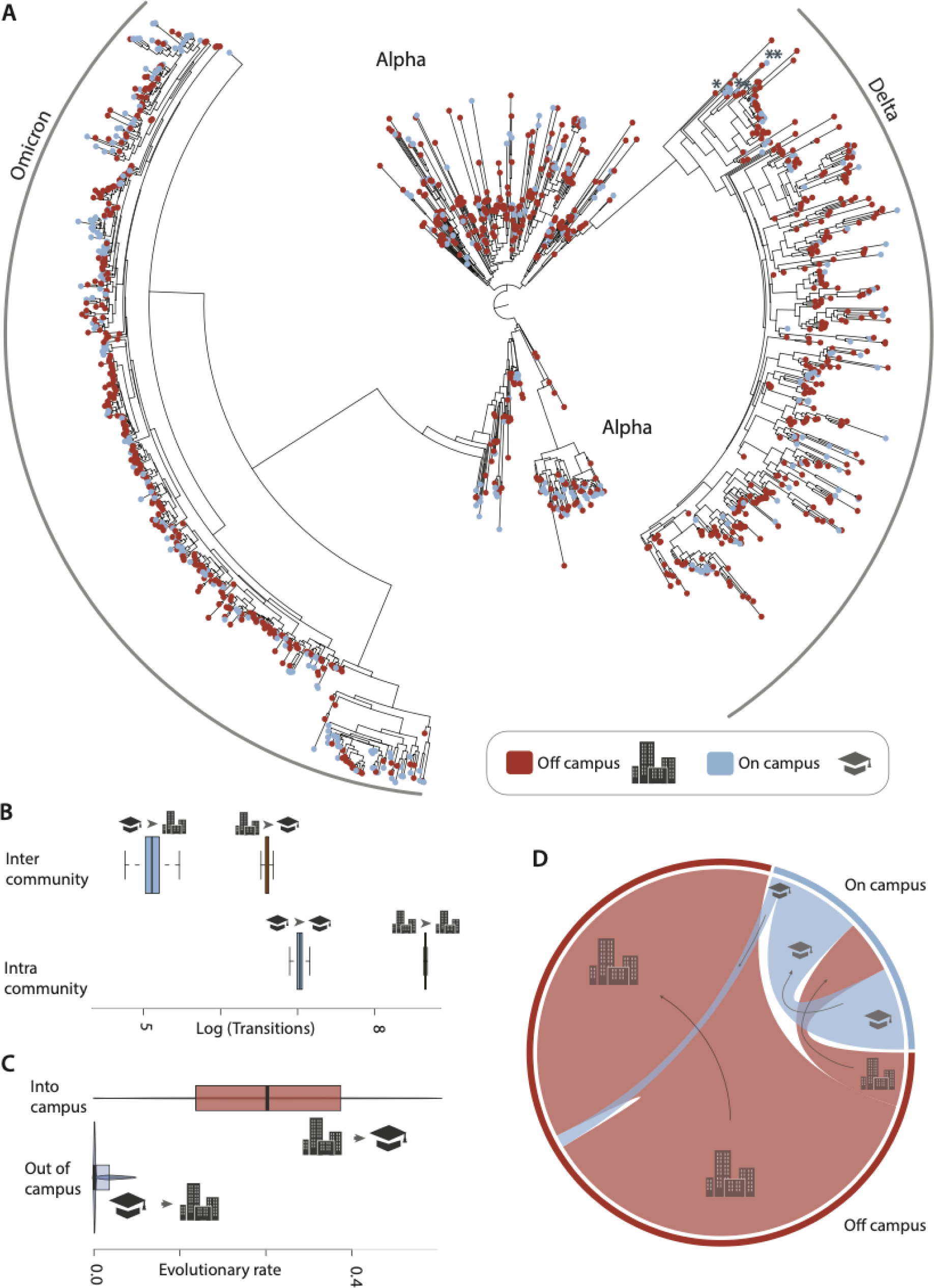
Transition history and transition dynamics between on and off campus communities during the study period. **(A)** Maximum likelihood tree topology of SARS-CoV-2 sequences collected during this study. Tip labels indicate whether samples were derived from on campus (cool shading) or off campus (warm shading) locations. * symbols in the delta strain portion of the phylogeny demarcate lineages identified as alpha by diagnostic mutations, but are phylogenetically resolved within delta (see supplemental materials). **(B)** Box plots depict the estimated quantiles of the log-transformed number of transitions occurring between communities. Top row reflects the frequency of transmissions occurring between the campus and off campus community (left) or between the off campus community and the campus (right). Bottom row reflects the frequency of transmissions within the campus (left) or within the off campus community (right). Cartoons above each box plot indicate the transmission mode and correspond to the legend in **(A)**. **(C)** The tempo of transmission between communities. Violin plots depict the estimated distribution of evolutionary transition rates estimated in BEAST, truncated to the 3rd quartile (75%). **(D)** Relative frequency of transmissions between and within communities. Chord diagram depicts the relative frequency of transmissions, shaded by community as in **(A)**. Outer bands correspond to total relative cases for each community, and inner chords illustrate transmission mode scaled to their frequency. Arrows and cartoons illustrate the direction of transmission.

Universities are often assumed to be amplifiers of viral transmission due to high-density living and frequent interpersonal interactions [11]. However, our results suggest that the off-campus student housing was the primary driver of infection trends at UNC Charlotte, with the on-campus population largely reflecting, rather than shaping, viral transmission dynamics among off-campus students. The clustering of on-campus infections following introductions from the off-campus population indicates that once introduced, transmission among those students living in the university environment was sustained at lower overall rates than among those living in the surrounding community. This pattern suggests that on-campus student residences are vulnerable to external seeding events and may not act as primary drivers of viral transmission in the community. As viral genomic data from COVID-19 pandemic continues to be analyzed, evidence for multiple introductions to urban college campuses that seeded in situ outbreaks continue to accrue, as does evidence for these environments generating only minimal spillover into their urban surroundings [21,33]. More broadly, these results suggest that on campus housing may follow similar transmission dynamics as other highly structured sub-populations such as nursing homes or long term care facilities [34]. It is becoming increasingly evident that community-wide mobility networks are predictive of infection pathways in the urban landscape [35], suggesting that the degree of connection of students to the community outside of campus is likely a major determinant of transmission into the campus. Our data lack fine-scale resolution concerning mobility between neighborhoods and individuals, precluding an analysis of whether certain areas have disproportionately seeded the campus. Regardless, our findings emphasize the importance of considering universities within the broader landscape of urban disease ecology [36,37]. Rather than viewing university dormitories as isolated entities shaping incidence patterns across a metropolitan region, integration of urban campus residence data into the broader urban community structure [38] will allow communities to harness information from universities’ healthcare platforms to understand how these settings are embedded within complex mobility networks that mediate pathogen flow across the urban landscape.

### Mitigation strategies impact transmission rates, not source-sink dynamics

To assess whether shifts in mitigation strategies corresponded to changes in transmission dynamics between on-campus and off-campus student housing, we analyzed SARS-CoV-2 lineage movement across four major phases of the pandemic: Wuhan (Fall 2020), Alpha (Spring–Summer 2021), Delta (Fall 2021), and Omicron (Spring 2022). Throughout this period, mitigation strategies transitioned from stringent controls—such as reducing dormitory occupancy to one-third capacity, enforcing masking and social distancing, and relocating SARS-CoV-2-positive students to designated isolation dormitories—to increasingly relaxed control measures that culminated in the resumption of in-person learning with many previous mitigation strategies optional (**see supplemental materials**). As measures relaxed, so did compliance with self-reported symptom tracking, mirroring broader trends in pandemic fatigue and shifting public health priorities [39–41]. These changes in campus policy largely aligned with evolving state and federal guidelines for North Carolina (**supplemental materials**), and raise the possibility that viral transmission dynamics between off and on campus may experience concomitant shifts that amplify spillover between these communities. However, our analyses strongly suggest this to not be the case.

Considering transmission dynamics across major variant-driven case surges of the pandemic reveals incidence trends that consistently aligned with mitigation shifts (**Figure 2**). In general, relaxation of restrictions led to higher frequencies of incidence in the on campus population, as well as a separation between the distribution of transitions between on campus cases, and transitions from off campus onto campus, with higher levels of on campus circulation (**Figure 2**). For example, during the initial Wuhan phase, when stringent mitigation measures such as mask mandates, social distancing, and routine testing were in place, on-campus transmission was equivalent to transitions from off to on campus (**Figure 2A**). In contrast, incidence levels during Alpha, when many restrictions were lifted, led to on campus transmission events outpacing the transitions from off to on campus (**Figure 2B**). This same pattern was repeated when comparing delta and omicron, the latter of which vastly exceeded transmission levels observed in other phases of the pandemic, both on and off campus (**Figure 2C&2D; Supplemental Table 1**). These incidence patterns reflect a global trend, as Omicron’s increased transmissibility and immune evasion properties contributed to surges of infection worldwide [42–44].

**Figure 2.**
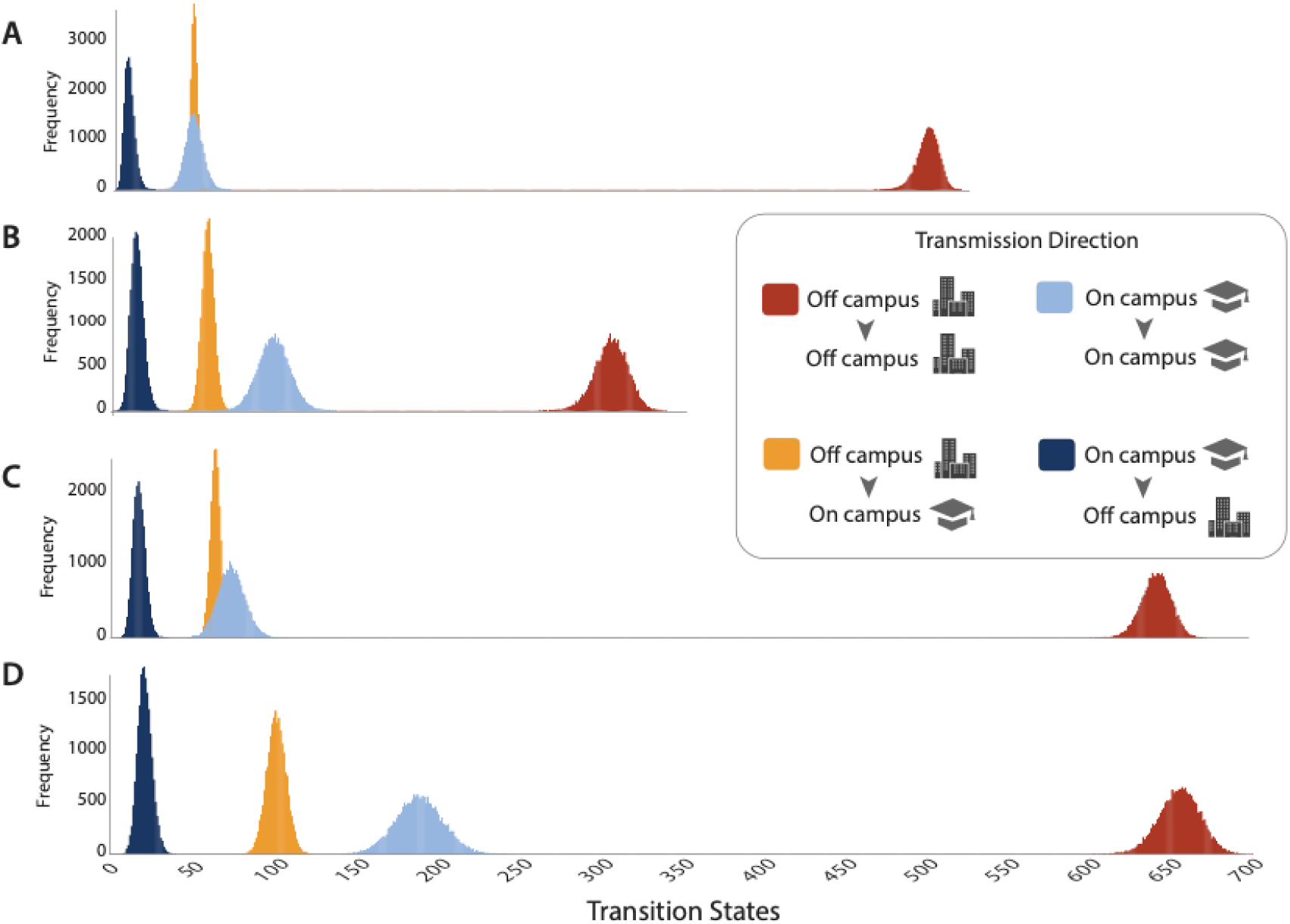
State transition distributions across four major waves of the pandemic: (A) Wuhan; (B) Alpha; (C) Delta; and (D) Omicron. The frequency (Y axis) of transitions (X axis) between and within residence types are indicated by the shadings in the figure legend with areas indicating the direction of transmission for each shading (i.e., off-campus to on-campus).

However, regardless of time period, movement of viruses from the campus residences into the off-campus community remained unchanged. Transmission occurred primarily from the off-campus residences into on-campus residences, rather than the reverse (**Figure 3**). Even as on-campus transmission increased during Omicron, there was no corresponding rise in outward viral movement from campus into the community (**Figure 2D**; **Figure 3**). These patterns were robust to resampling, indicating this result is not an artifact of differential incidence levels between on-campus and off-campus populations alone (**Supplemental Figure 2**). Instead, the persistent asymmetry between off-campus to on-campus and on-campus to off-campus transitions suggests that the primary drivers of viral introduction into the university were external, rather than originating within campus populations. This pattern mirrors findings from other university settings during Omicron, where campuses exhibited high in situ transmission but limited spillover into the surrounding community [9,45]. These results suggest that, regardless of mitigation measures, UNC Charlotte continued to act primarily as a sink rather than a source of community transmission.

**Figure 3.**
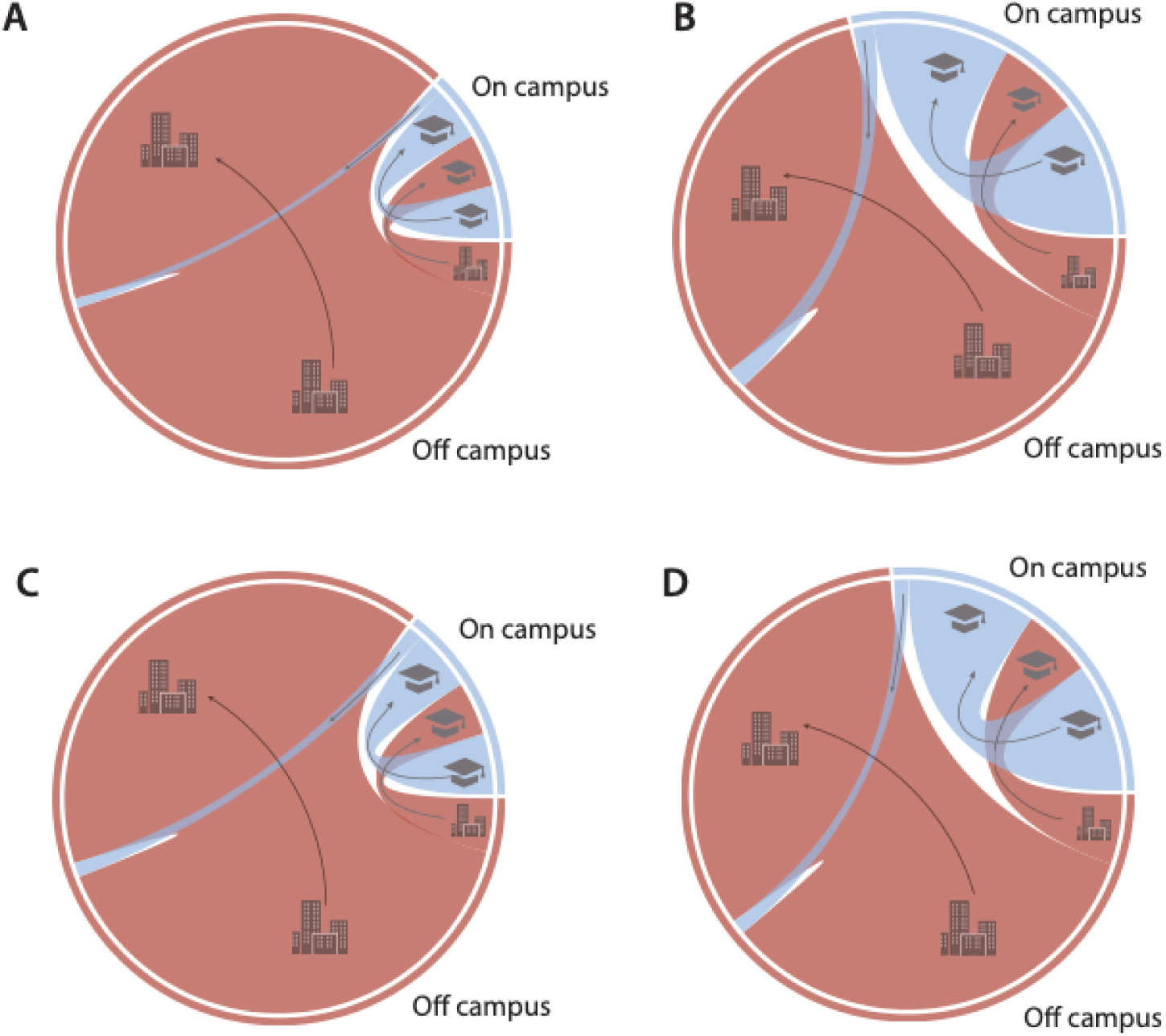
Transmission dynamics between on and off campus across variant waves: (A) Wuhan; (B) Alpha; (C) Delta; and (D) Omicron. The relative frequency of transmissions between and within communities is depicted in chord diagram, shaded by community as in Figure 1. Outer bands correspond to total relative cases for each community, and inner chords illustrate transmission mode scaled to their frequency. Arrows and cartoons illustrate the direction of transmission.

There is no doubt that initial pandemic restrictions and mitigation strategies saved millions of lives across the globe [46]. Similarly, targeted university interventions such as masking mandates and routine testing are estimated to have reduced infections on University campuses by an order of magnitude or more [47]. However, universities, like other sectors [48], had to navigate a complex trade-off between the immediate health benefits of mitigation and the long-term academic, social, and economic consequences of restrictive policies [49,50]. Advances in genomic surveillance methods [51,52] as well as computational models have come to play a crucial role in balancing disease suppression with the need for in-person education [53,54]. Our results suggest that placing the university within the broader framework of urban disease ecology could provide vital additional context to multiple urban sectors. In particular, our finding of a marked asymmetry in viral transmission, that persisted across mitigation strategies, suggests that on campus residences such as those at UNC Charlotte may be epidemiologically insular, acting as relatively self-contained transmission units rather than perpetual community wide spreaders. Further work leveraging computational [55] and phylodynamic approaches [26,56] are critically needed to assess whether similar trends hold across other urban campuses.

### On campus infection waves lag behind off campus waves

If on-campus student housing were a major source of viral transmission, we would expect infection waves among dormitory residents to precede or at least coincide with those in off-campus housing. However, our phylodynamic analyses reveal a consistent lag in on-campus infection waves relative to those in off-campus housing, with peaks in on-campus lineage counts occurring after surges in off-campus populations (**Figure 4A**). This pattern is particularly pronounced for the Delta and Omicron waves (**Figure 4**), where variant-driven outbreaks first became established in off-campus housing before spreading into dormitories. Incidence peaks largely align with the broader emergence of new variants of concern in the surrounding community (**Figure 4B**), and the delayed pattern of spread is consistent with the expected transmission lag when considering the average incubation period of SARS-CoV-2, which ranges from 5 to 14 days [57,58]. The variation between off and on campus incidence patterns over time was particularly pronounced during periods of higher mitigation on campus, which resulted in an overall muting of peak incidence patterns (**Supplemental Figure 3**), and likely reflects the efficacy of mitigation measures. Collectively, these results imply that consistent transmission from source off-campus residences seeded numerous on campus cases and outbreaks during all waves of the pandemic, with times of heightened use of on-campus mitigation strategies, controlled environments, and testing preventing further in situ outbreaks.

**Figure 4.**
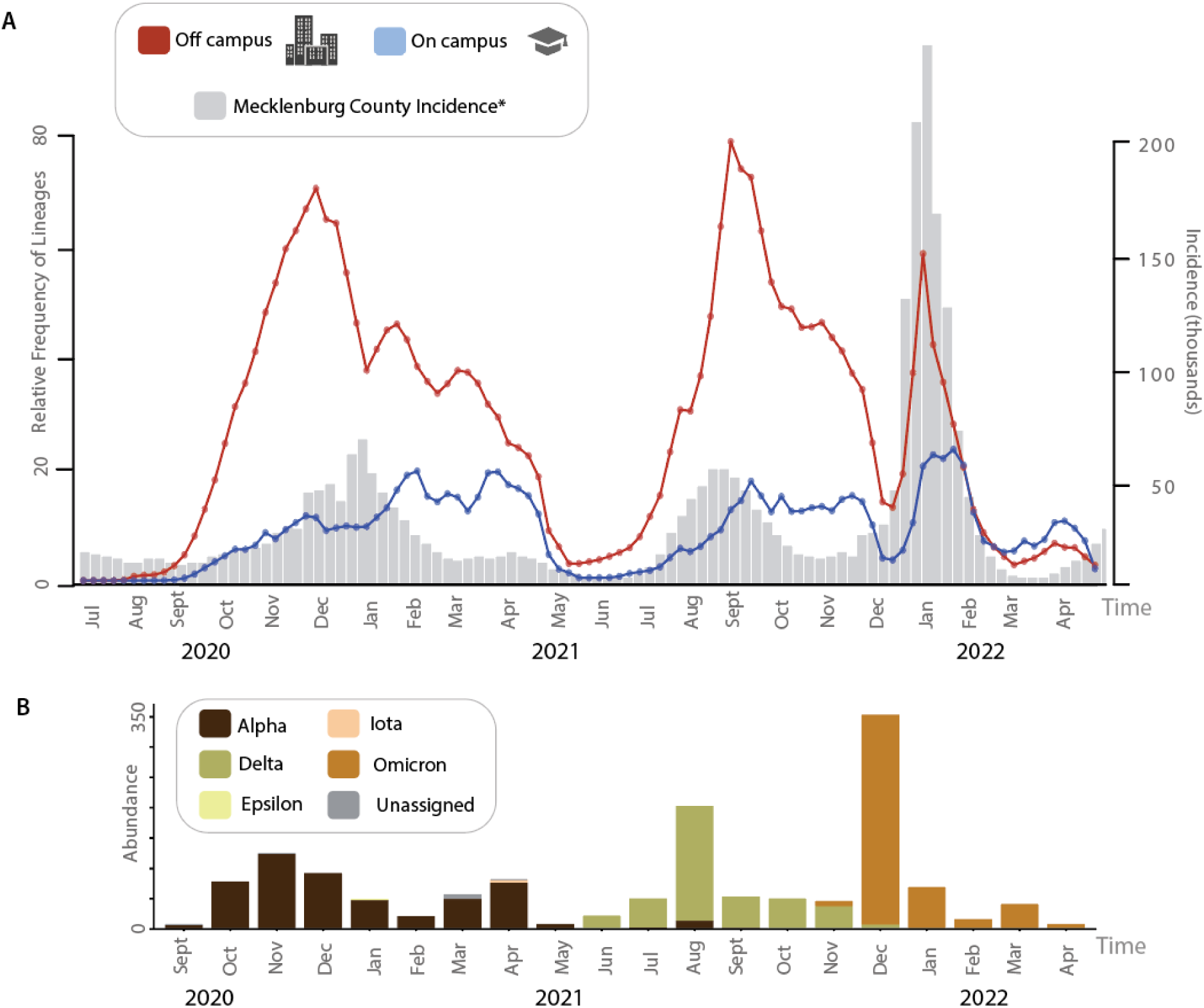
Trends in lineages through time and incidence. (A) SARS-COV-2 relative frequency of lineages through time for off-campus (warm shading) and on-campus (cool shading) residences plotted in comparison to incidence in Mecklenburg county (gray). Circles in lineage through time plots represent specific time points. **(B)**. Abundance of circulating strains during the time period depicted in **A**. Strains are indicated in the legend.

Throughout the COVID-19 pandemic, transmission dynamics often varied spatially, with some areas experiencing infection waves weeks ahead of others [59]. This staggered pattern of viral spread can occur both at larger geographic scales (e.g., hemisphere, country, etc) as well as at more localized scales (e.g., state, county, etc) [60,61]. Understanding variation in the spread of disease is of particular concern in densely populated urban environments where social connectivity, mobility patterns, and mitigation strategies influence the timing and intensity of outbreaks [62]. If universities were major sources of community spread, we would expect campus surges to precede infection waves in off-campus residences, that, in turn, would seed transmission through the larger metropolitan area. However, our findings strongly suggest the reverse. The on campus residences at UNC Charlotte largely acted as recipient of community viral spread, rather than an independent epicenter of transmission. This pattern of transmission raises the possibility that off campus housing acts as a gateway enabling viruses circulating in the community to enter and circulate on campus. Whether there are asymmetrical pathways of transmission such as this between the broader community and the campus remains unknown and represents an exciting area of future research.

## Conclusion

Our findings demonstrate that on-campus student housing at UNC Charlotte functioned as a transmission sink rather than a source of community-wide viral spread throughout the COVID-19 pandemic. By leveraging whole-genome sequencing and Bayesian phylogenetic analyses, we reconstructed SARS-CoV-2 transmission pathways and found strong evidence that viral introductions into dormitories overwhelmingly originated from off-campus housing rather than spreading outward from campus into the broader student population. This pattern persisted across multiple pandemic waves and mitigation phases, suggesting a consistent epidemiological role for on-campus residences as recipients of community transmission rather than amplifiers of broader outbreaks. Importantly, even during periods of high incidence—such as the Omicron surge—there was no corresponding increase in outward viral movement from dormitories to off-campus residences. These results suggest on-campus housing to possibly be similar in expected viral transmission dynamics to other structured, high-density subpopulations that have limited interactions with the broader community, such as long-term care or military facilities, in which external introductions predominantly seed internal outbreaks [63–65]. Future work assessing additional universities with similar on and off campus residence structures is critically needed to determine the degree to which these transmission source-sink dynamics persist across institutional and geographic contexts.

## Materials and Methods

### Sample Collection and RNA extraction

Samples were obtained from the Covid Testing Center (later absorbed into the Student Health Center) at the University of North Carolina Charlotte, and StarMed, an urgent care network serving Mecklenburg and surrounding counties. Collection involved buccal nasal swabs from symptomatic as well as asymptomatic individuals, followed by viral RNA extraction by the respective facility. StarMed utilized the Kingfisher system with the MagMAX™-96 Total RNA Isolation Kit (Catalog No: AM1830) for RNA extraction, while the UNC Charlotte testing center employed the KingFisher Flex with the MagMAX™ Viral/Pathogen II (MVP II) Nucleic Acid Isolation Kit (Catalog No: A48383). After extraction, qPCR was conducted to detect SARS-CoV-2 positive samples. UNC Charlotte used three probes (N1, N2, and S primers, designed by the CDC) with the TaqPath™ COVID-19 Combo Kit (Catalog No: A47814), whereas StarMed relied solely on the N1 primer for detection, using the same kit. Only RNA from SARS-CoV-2 positive samples was forwarded for sequencing. This sampling resulted in 1444 unique positive cases of which 1431 were successfully sequenced. This sample represents nearly 90% of all known campus associated SARS-CoV-2 cases during the study period, with only approximately 200 additional cases voluntarily reported by individuals who were tested in locations away from campus without a surveillance sequencing program.

### Library preparation and sequencing

RNA extracts of clinical samples were reverse-transcribed and amplified in PCR according to a tiling amplicon protocol modified from Pater et al. [66]. A fully detailed guide to this modified protocol is available on protocols.io [67]. Briefly, first we transformed extracted RNA into complementary DNA (cDNA), with cDNA enrichment of the SARS-CoV-2 genome performed by multiplexing PCR with two pools of ARTIC primers. For each sample, the ARTIC PCR products were pooled and amplicons were isolated through magnetic Solid Phase Reversible Immobilization (SPRI) beads and quantified by Qubit fluorescence. Subsequent sequencing library preparation comprised amplicon end-prep, sample barcoding, sample pooling and cleaning, and sequencing adapter ligation. Finally, the sample library was loaded onto an R9 flow cell (Oxford Nanopore Technologies, ONT) and sequenced using the ONT PromethION instrument. Manufacturer-recommended settings for the LSK-109 genomic sequencing by ligation kit, Native Barcode Expansion Kit (Exp-NBD196), and FLO-PRO002 flow cell were applied at the start of the run.

### Sequence assembly and variant calling

Basecalling was initially carried out in real time using the Oxford Nanopore guppy software with high accuracy setting, via the MinKnow user interface on the ONT PromethION instrument. During spring of 2021, updates to ONT basecalling algorithms became available and were applied as they were released. In order to avoid biases arising due to changes in basecalling algorithms, sequences were re-basecalled from raw fast5 files using the ONT 1.2.1 “super high accuracy” basecalling method. Assembly and variant calling of SARS-CoV-2 sequences was performed using a modified version of the ARTIC ‘fieldbioinformatics’ pipeline (https://github.com/artic-network/fieldbioinformatics). Briefly, reads were filtered and trimmed with guppyplex to remove V3 adapter sequences and to exclude chimeric sequences smaller than 250 bases and longer than 700 bases. Alignments were constructed using minimap2, followed by medaka (https://github.com/nanoporetech/medaka) for polishing, variant calling, and consensus building using the ‘r941_prom_high_g360’ model.

Consensus sequences were classified using Pangolin (https://github.com/cov-lineages/pangolin) versions 3.1.11, 3.1.14, 3.1.16, 3.1.17, 3.1.19, 3.1.20, and 4.1.2 to ensure accurate lineage assignments over the course of the study (For example, earlier versions would not be able to accurately classify Delta or Omicron due to their unique mutation signature). Sequences were inspected for quality based on the pass/fail metrics in Pangolin that assess minimum sequence length and maximum allowable ambiguous bases (N content).. Of the sequenceable samples, 13 failed quality control at this classification step and were excluded from downstream analyses. Additional quality control was conducted using Nextclade [68], which assesses missing data, ambiguous sites, mutation clusters, stop codons, and frame shifts for SARS-CoV-2 (https://github.com/nextstrain/ncov).

### Phylogenetic Analyses

A total of 1444 viral samples were collected from both on-campus and off-campus populations between September, 2020 and May, 2022. Of these, 1431 passed the sequence quality control assessments described above. The final set of sequences (available in GISAID with EpiSet ID: EPI_SET_250220my) is composed of 470 samples originating from on-campus students and 961 from off-campus individuals. We used the Wuhan strain (accession NC_045512) to serve as a reference genome and outgroup for analysis. Sequences were aligned using MAFFT v7.525 [69]. To estimate the evolutionary history of our collected samples, we used MAPLE (MAximum Parsimonious Likelihood Likelihood Estimation), a computational tool designed for large-scale phylogenetic inference of epidemiological datasets that has been shown to significantly reduce computational analysis time while increasing accuracy relative to other likelihood approaches [70].

We used RelTime with Dated Tips (RTDT) [71] in Mega X [72] to time calibrate the MAPLE inferred maximum likelihood topology, allowing us to estimate the transmission timing of SARS-CoV-2 across on-campus and off-campus student housing. RTDT is an algebraic method designed for estimating divergence times from temporally sampled molecular sequences, making it particularly well-suited for analyzing pandemic-scale datasets with precise sampling time information. RTDT incorporates a relative rate framework, enabling computational efficiency while maintaining a level of accuracy that is par with more computationally complex Bayesian approaches to divergence time estimation such as those implemented in BEAST [73–75], including for the analysis of SARS-CoV-2 data [76]. Divergence times were calibrated using the time each viral genome was sampled.

### Reconstructing the history of transmission

We used the discrete phylogeographic diffusion model implemented in BEAST v2.6.7 [32,77,78] to reconstruct the transmission history of SARS-CoV-2 between student housing communities. This model integrates ancestral state reconstruction (ASR), a well-established method in evolutionary inference [79–82], with a diffusion model that models viral spread through distinct transmission events between predefined geographic units [83]. This approach allowed us to quantify the directionality and frequency of transitions between on-campus and off-campus housing while integrating branch-specific rate variation to accommodate heterogeneity in transmission dynamics [83]. We fixed the time calibrated phylogeny estimated above, and ran two independent markov chains for up to 250,000,000 generations to estimate transmission histories. Runs were stopped at 232,979,000 generations as inspection of the likelihood and other parameter values in Tracer v1.7.2 [84] indicated convergence between runs, with all effective sample size (ESS) values above 200, indicating effective sampling of the target distributions. To verify that posterior estimates were informed by the empirical data, we conducted an additional BEAST analysis in which parameters were sampled exclusively from the prior distributions.. We used the Babel package (https://github.com/rbouckaert/Babel) to quantify the distribution of on or off campus lineages through time with the LineagesThroughTimeCounter function, and also quantified transitions between on and off campus using the transitionStateCounter function. Resulting transition histories were imported into R visualized using Circos plots with functions from circlize v0.4.16 [85,86] treedataverse v0.0.1 [87,88], and tidyverse v0.4.6 [89] libraries for additional visualizations. Transmission histories were visually compared to incidence frequencies in Mecklenburg County using public incidence data available from the North Carolina Respiratory Virus Summary Dashboard (https://covid19.ncdhhs.gov/dashboard).

Our dataset captured approximately 90% of all cases known within these populations during the study period. However, it is possible that the uneven levels of incidence between off and on campus might bias estimation of transmission dynamics. To assess the degree to which possible sampling biases of campus populations impact estimated transmission dynamics, the above analyses were repeated on randomly subsampled data sets with predefined on-campus:off-campus ratios (1:5, 2:3, 1:1, 3:2, 5:1). For each set of datasets at a given ratio (e.g. 5 on campus sampler per very 1 off campus sample), total transmissions between and within population (off-campus to off-campus, on-campus to on-campus, off-campus to on-campus, and on-campus to off-campus) were quantified to ensure a consistent signal of source-sink dynamics relative to that based on the observed incidences of COVID-19.

## Data Availability

All data produced in the study are available as a GISAID EpiSet or as part of a Zenodo data package. DOI/URL information for supplementary materials shared via EpiCoV, Zenodo and GitHub are provided.

https://doi.org/10.55876/gis8.250220my

https://doi.org/10.5281/zenodo.14946855

https://github.com/jbolanos93/Covid-Campus-Phylodynamics.git

## Acknowledgements

Dr. Robert Jones at the UNC Charlotte Student Health Center and Dr. Angelica Martins in the UNC Charlotte Division of Research provided samples and COVID-19 diagnostic test metadata as specified under UNC Charlotte protocol IRB 21-0113. We thank them, their teams, and the other staff of the UNC Charlotte Division of Research for their collaboration and tireless public health efforts during the COVID-19 pandemic emergency.

## Declaration of generative AI and AI-assisted technologies in the writing process

During the preparation of this work the author(s) used ChatGPT in order to obtain feedback and enhance the manuscript. After using this tool/service, the author(s) reviewed and edited the content as needed and take(s) full responsibility for the content of the publication.

## CRediT author roles

Bolanos: Conceptualization, Methodology, Investigation, Formal Analysis, Visualization, Software, Writing – Original Draft, Writing – Review and Edit

Dornburg: Conceptualization, Methodology, Visualization, Funding Acquisition, Writing – Original Draft, Writing – Review and Edit

Harris: Resources, Data Curation, Investigation

Kunkleman: Data Curation, Software, Methodology

Ferdous: Methodology, Investigation

Taylor: Project Administration, Methodology, Investigation

Schlueter: Funding Acquisition, Project Administration, Supervision, Writing – Review and Edit

Gibas: Conceptualization, Funding Acquisition, Project Administration, Supervision, Visualization, Writing – Original Draft, Writing – Review and Edit

## Data Availability

SARS-CoV-2 sequence has been deposited to GISAID (doi: 10.55876/gis8.250220my). All code used for analyses has been archived on Zenodo (DOI:10.5281/zenodo.14946855; URL: https://doi.org/10.5281/zenodo.14946855).

