## Supplemental Materials for "On-Campus Dormitories as Viral Transmission Sinks: Phylodynamic Insights into Student Housing Networks During the COVID-19 Pandemic"

**Supplemental Materials For:**
**Viral Dispersal Patterns in University Communities: analysis of comprehensive COVID-19 sequencing on an urban university campus.**

Juan Bolanos, Alex Dornburg, April Harris, Samuel Kunkleman, Jannatul Ferdous Moon, William Taylor, Jessica Schlueter, Cynthia Gibas

**Supplemental Results… … … … … … … … … … … … … … … … … .. 2**

**Supplemental Figures… … … … … … … … … … … … … … … … … .. 3-5**

**Supplemental Table … … … … … … … … … … … … … … … … … … .6**

**References… …. …. … … … … … … … … … … … … … … … … … .7**

### Supplemental Results

*COVID-19 intervention strategies shifted through time at UNCC*

Throughout the study period, the University of North Carolina at Charlotte (UNCC) implemented a multi-layered mitigation strategy, including thrice-weekly wastewater surveillance at the building and neighborhood levels, with follow-up clinical testing for positive wastewater signals [1]. Following the onset of the COVID-19 pandemic, UNCC resumed in-person operations with reduced occupancy in on-campus housing in September 2020. Throughout the 2020-21 academic year, dormitory populations were reduced to one-third of typical capacity, masking and social distancing were required, and students testing positive for SARS-CoV-2 were relocated to a designated isolation dormitory. Additional measures implemented during this included symptomatic testing, mandatory asymptomatic testing before the start of each semester, an in-house contact tracing program, and voluntary self-reporting of symptoms through a daily survey. These extensive efforts earned UNCC the AASCU Excellence and Innovation Award for Campus Pandemic Response [2]. However, campus COVID-19 mitigation strategies evolved in response to shifting pandemic conditions (**Supplemental Figure 1**). Over the course of the 2020-2021 academic year, mitigation measures were gradually relaxed, and adherence to daily symptom reporting via health surveys steadily declined. By August of 2021, dormitories were reopened at full capacity. Following the return to campus, the emergence of the delta variant catalyzed a reinstatement of prior mitigation strategies, including masking and social distancing strategies as well as building-wide testing after detection of virus in building wastewater and increased re-entry and random asymptomatic testing. This was combined with incentivization of vaccine adoption, by allowing exemption from routine individual testing. Throughout the course of the fall semester, these measures again relaxed. By the time of the peak omicron wave in January 2022, in-person learning resumed with significantly reduced mitigation protocols, rendering the majority of previous strategies optional. This shift marked a transition from structured, enforced measures to a reliance on personal responsibility, mirroring broader trends in institutional pandemic responses reflecting changing public health guidance [3] as well as shifts in the immune status of the general population following infection or vaccination [4,5].

### Supplemental Figures

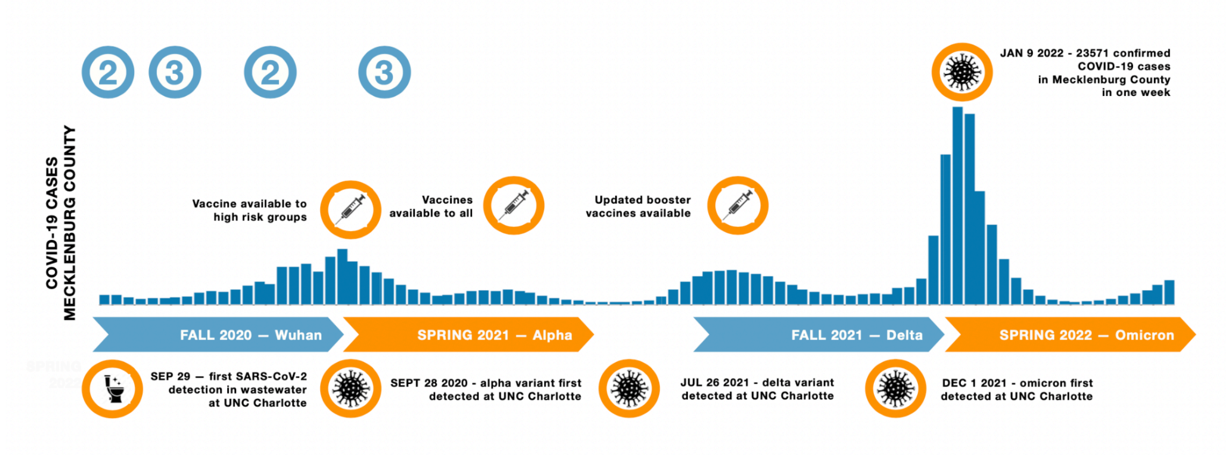


**Supplemental Figure 1. Timeline of major COVID-19 pandemic events at UNC Charlotte and in Mecklenburg County, NC, relative to NC-DHHS case incidence data for the study period (August 15, 2020 to May 15, 2022)**. We divided the timeline into four phases aligned to university semesters. The later time periods roughly correspond to the timing of introduction to North Carolina of the dominant variants of concern (VOCs) that emerged during winter 2020 (alpha), summer 2021 (delta) and winter 2021 (omicron) and are named accordingly. Reported COVID-19 incidence is depicted by the cool bar graph over the course of the study. Vaccination rollouts and other events are indicated by warm circles at their corresponding time points. Cool circles labelled (2) or (3) correspond to the emergency conditions designated by the state. North Carolina went into full lock down–Phase 1 conditions– at the onset of the COVID-19 pandemic. However, during the period of this study the state of North Carolina had lifted phase 1 conditions, remaining primarily in Phase 2 or Phase 3 of emergency guidelines. UNC Charlotte mirrored these guidelines. The campus remained open during the entire period, managing exposure via density reduction between August 2020 and May 2021, and returning to full occupancy for the second year of the pandemic declaration. The University's actions in the first year closely paralleled the statewide phases. In the second year, the university delayed the start of in-person instruction each semester by 2-3 weeks, due to the delta and omicron waves. During the entire 2-year period, wastewater surveillance of dormitories triggered testing of on-campus residents, and additional re-entry, symptomatic, case contact, and random surveillance testing of the student population continued during that time (see supplemental materials for additional details).

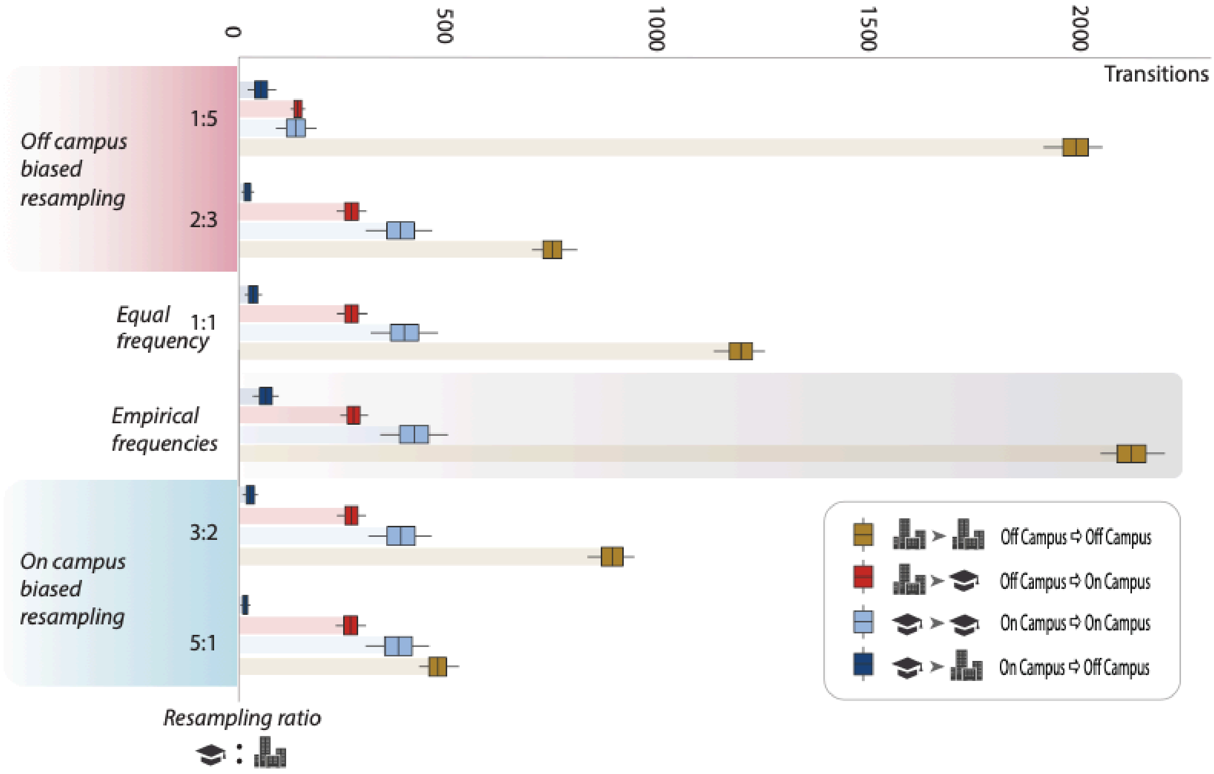


**Supplemental Figure 2: Effect of Subsampling on Estimated Transmission Dynamics.** State transition frequencies were evaluated across phylogenies randomly subsampled at predefined on-campus:off-campus sampling ratios (1:5, 2:3, 1:1, 3:2, 5:1). Transition categories (off-campus to off-campus, on-campus to on-campus, off-campus to on-campus, and on-campus to off-campus) were compared across biased sampling strategies to assess if uneven frequencies of infection between populations would impact the trends observed in the empirical frequencies. Across all sampling strategies, the relative relationship between transition frequencies remained consistent, with only the absolute number of transmission events within the off-campus community modulating. Bars represent the mean number of transitions across each simulation and category, with box plots indicating the 95% confidence interval for each distribution. The x-axis denotes the resampling ratio, and the y-axis indicates the number of inferred transmission events. Off campus and on campus biased resampling results are indicated by shading on the x-axis.

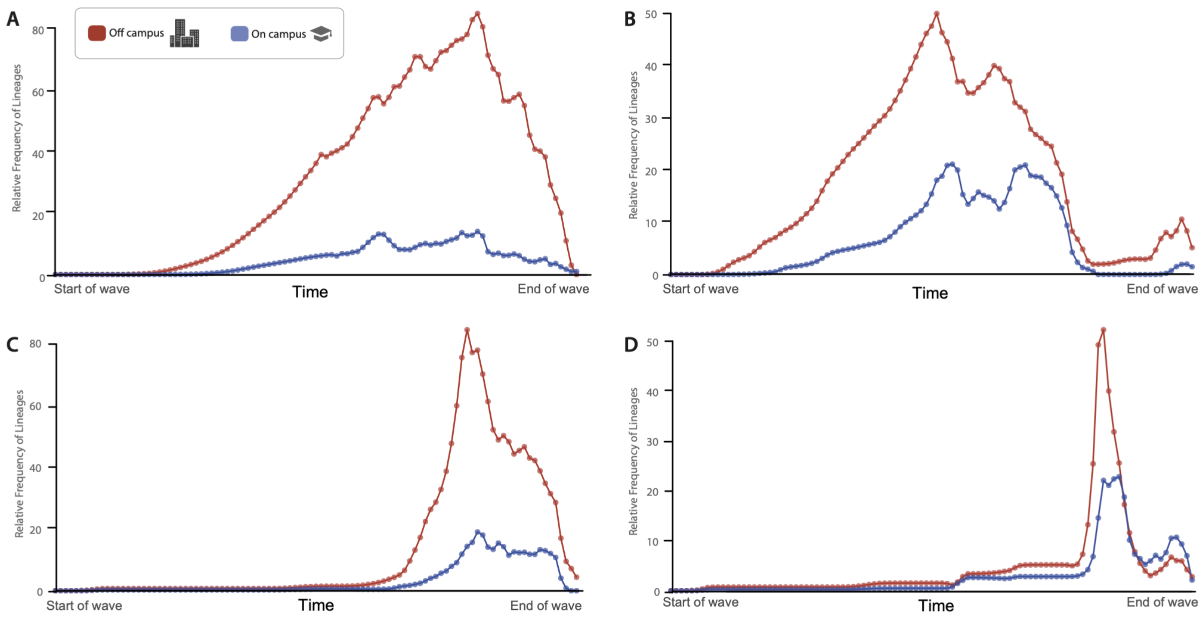


**Supplemental Figure 3**. **Trends in lineages through time and incidence during the major waves of COVID-19 pandemic that occurred during the study period (A) Wuhan, (B) Alpha, (C) Delta, (D) Omicron.** Lines indicate the relative frequency of SARS-COV-2 lineages through time for off-campus (warm shading) and on-campus (cool shading). Circles in lineage through time plots represent specific time points.

**Supplemental Table**

**Supplemental Table 1: Mean number of transmissions estimated within and between populations during major waves of the COVID-19 Pandemic**

| Wave | From | To | Transitions |
| --- | --- | --- | --- |
| **Wuhan** | Off Campus | Off Campus | 500 |
| | Off Campus | On Campus | 48 |
| | On Campus | Off Campus | 8 |
| | On Campus | On Campus | 48 |
| **Alpha** | Off Campus | Off Campus | 305 |
| | Off Campus | On Campus | 58 |
| | On Campus | Off Campus | 15 |
| | On Campus | On Campus | 98 |
| **Delta** | Off Campus | Off Campus | 644 |
| | Off Campus | On Campus | 64 |
| | On Campus | Off Campus | 17 |
| | On Campus | On Campus | 74 |
| **Omicron** | Off Campus | Off Campus | 658 |
| | Off Campus | On Campus | 102 |
| | On Campus | Off Campus | 21 |
| | On Campus | On Campus | 190 |
